# Effectiveness of Mulligan joint mobilizations and trunk stabilization exercises versus isometric knee strengthening in the management of knee osteoarthritis: study protocol for a randomized controlled trial

**DOI:** 10.1101/2020.05.03.20089276

**Authors:** Shaikh Nabi Bukhsh Nazir, Syed Shahzad Ali, Saeed Akhtar

**Author notes:** ***Correspondence:*** Dr. Shaikh Nabi Bukhsh Nazir (PT), Department of Physiotherapy, Institute of Physical Medicine & Rehabilitation, Dow University of Health Sciences, Karachi, Pakistan.

## Abstract

**Background:** Knee Osteoarthritis (KOA) has a huge negative impact on gait parameters and on many components of biomechanics, including impairment of dynamic lower limb alignment and control of lumbopelvic movement. Specifically addressing these problems in treatment regimens is therefore critical but they must first be studied in detail before they can be used clinically to treat patients with KOA. This study protocol focuses on whether Mulligan joint mobilization with movement demonstrably decreases pain and restores healthy joint biomechanics and whether trunk stabilization exercises improve stability of the trunk, thereby distributing the weight of the body evenly on both feet. Because the treatment effects of neither treatment are conclusive, this study aims to determine their efficacy versus isometric knee strengthening.

**Method:** The study protocol is a three-arm randomized controlled trial. After initial screening by a referring consultant, subjects who fulfil the study criteria will be randomly assigned to one of three groups. They will then be given an explanation of study objective and asked for their informed consent to participate in the study. Group 1 will receive Mulligan mobilization with kinesiotaping and knee strengthening. Group 2 will receive trunk stabilization exercises, knee strengthening, and kinesiotaping. Group 3 will receive knee strengthening along with kinesiotaping. All participants will be evaluated using a visual analogue scale, Knee injury and Osteoarthritis Outcome Score, stair climb test and 6-minute walk test at the baseline, 3rd and end of 6th week.

**Discussion:** The results of this study will answer focused questions concerning the relative efficacy of each treatment in KOA patients. The findings of this study will inform clinical decision-making by healthcare professionals and researchers.

**Trial registration:** NCT04099017

## Background

Walking difficulty is commonly experienced by individuals with knee osteoarthritis (KOA), restricting their ability to participate in activities of daily living (ADLs). The prevalence of walking difficulty in KOA was found to be 30%, which correlates not only with an increased institutionalization rate and health care cost, but also with decreased quality of life. KOA is also associated with metabolic and systemic diseases such as hypercholesterolemia, high blood glucose levels, and high blood pressure. In turn, these complications can lead to even more serious health issues, thereby further increasing the economic burden. There is therefore a need to devise a cost effective treatment strategy to limit functional decline, joint pain and stiffness.^1^

People with osteoarthritis tend to avoid activity due to severe pain, but exercise has proved to be an effective treatment for this condition, reducing pain and improving physical function by improving the strength of the muscles surrounding the knee joint. Twenty-four sessions of therapeutic exercise appear to provide the most beneficial effects, but these effects have not been tested for walking capacity in a 6-minute walk test.^2^ However, multimodal exercise programs for the knee are known to provide better pain relief than knee strengthening alone.^3–4^

Weakness around the trunk muscles plays a crucial role in the development of knee pain, which is also linked with decreased strength of the trunk side flexor, hip abductor, lateral rotator and extensor muscles.^5–8^ However, there is limited evidence to suggest that trunk stabilization combined with either knee strengthening or trunk stability exercises improves lumbopelvic control and walking capacity. ^9–10^

Several studies have revealed the positive effects of joint mobilization with exercise in the management of KOA.^11–12^ The American College of Rheumatology (ACR) also recommends that patients with knee OA receive manual physical therapy in combination with knee strengthening exercises under the supervision of a qualified physiotherapist.^13^

In 1980, Brian Mulligan proposed a joint mobilization technique for the management of various musculoskeletal conditions to increase movement and reduce joint pain. Mulligan joint mobilization includes active movement and mobilization of the joint, which is termed Mobilization with Movement (MWM). MWM works on the principle of restoring the biomechanics of the knee by overcoming positional fault.^14–17^ A recent systematic review highlighted the problem that MWM only exhibits the immediate effects on pain and disability. The long-term effectiveness of MWM and well-designed a RCT have yet to be studied.^18^ It is known, however, that taping accompanied with the mobilization helps to support the correcting effects of mobilization and contributes to further reduction in the intensity of pain and the level of disability.^19–20^

Because the efficacy of KT in the management of KOA remains uncertain. A well-designed RCT is needed to evaluate the effects of elastic taping on pain, disability and sub-maximal exercise performance, especially in combination with exercise.^21^

At present, there are no RCT studies of the effects of Mulligan joint mobilization during 24 supervised exercise sessions. There are also few high-quality methodological studies investigating the effects of Mulligan mobilization on the management of KOA; especially the 6-minute walk test (walking difficulty), pain and disability. Therefore, we designed a randomized controlled trial to determine whether Mulligan joint mobilizations are better than trunk stabilization and isometric knee strengthening, at providing pain relief, reducing disability and improving sub-maximal exercise performance level in the KOA.

## Methods

### Study design

The three-arm randomized controlled trial will be carried out at the Institute of Physical Medicine and Rehabilitation, Dow University of Health Sciences. This study has been approved by the Institutional Review Board (IRB), Dow University of Health Sciences (IRB-1433/DUHS/Approval/2019/118). The randomized controlled study will follow SPIRIT, CONSORT and Tidier guidelines.

### Study population and allocation

A physiatrist and an orthopedic surgeon will perform clinical examinations of participants and evaluate x-rays taken within a three month period. Those participants who fulfil the inclusion criteria will be selected from the Outpatient Department of the Institute of Physical Medicine and Rehabilitation at Dow University of Health Sciences and the Orthopaedic Department of the Civil Hospital, both in Karachi. All patients will participate voluntarily and be required to sign an informed consent form after receiving oral and written explanations of the purpose of the study and the procedures to be used.

### Inclusion criteria

- Men and women aged 40 – 60 years.
- Knee osteoarthritis Grade I and II based on Kellgren and Lawrence (K/L) radiological criteria.^22^
- Confirmed diagnosis of KOA based on the clinical criteria of ACR.^23^
- Use of medicines such as vitamin D and calcium supplements

### Exclusion criteria

- Known skin allergies to Kinesiotaping
- Sensory-motor dysfunction of lower extremity
- Severe joint deformity of lower extremity
- Post Traumatic Arthritis
- Constitutional symptoms (fever, malaise, weight loss and high blood pressure)
- lower back pain
- History of spinal surgery
- Using assistive devices for ambulation, i.e. cane, walkers, sticks
- Receipt of physiotherapy treatment in the past three months
- Visual Analogue Scale <4
- Patello-femoral joint arthritis

### Randomization and allocation concealment

The study subjects will be randomly assigned to one of three groups (ratio 1:1:1) using a research randomizer (www.random.org). The schedule will be concealed using sequentially numbered opaque, sealed envelopes. The envelopes will be stored in a locker and opened in sequence within each stratum to reveal group allocation.

### Masking

After confirming the eligibility of the subjects, a physiotherapist will perform interventions for each group after obtaining treatment assignment from S.S. Ali. The subjects will not be aware of the group to which they are assigned, and different time slots will be given for each intervention. Another physiotherapist. who will also not be aware of allocation concealment, will record the reading of outcomes at the baseline, after three and six weeks.

### Interventions

The intervention frequency will be four sessions a week on alternate days for 6 weeks, i.e. 24 sessions in all.

**Figure 1:**
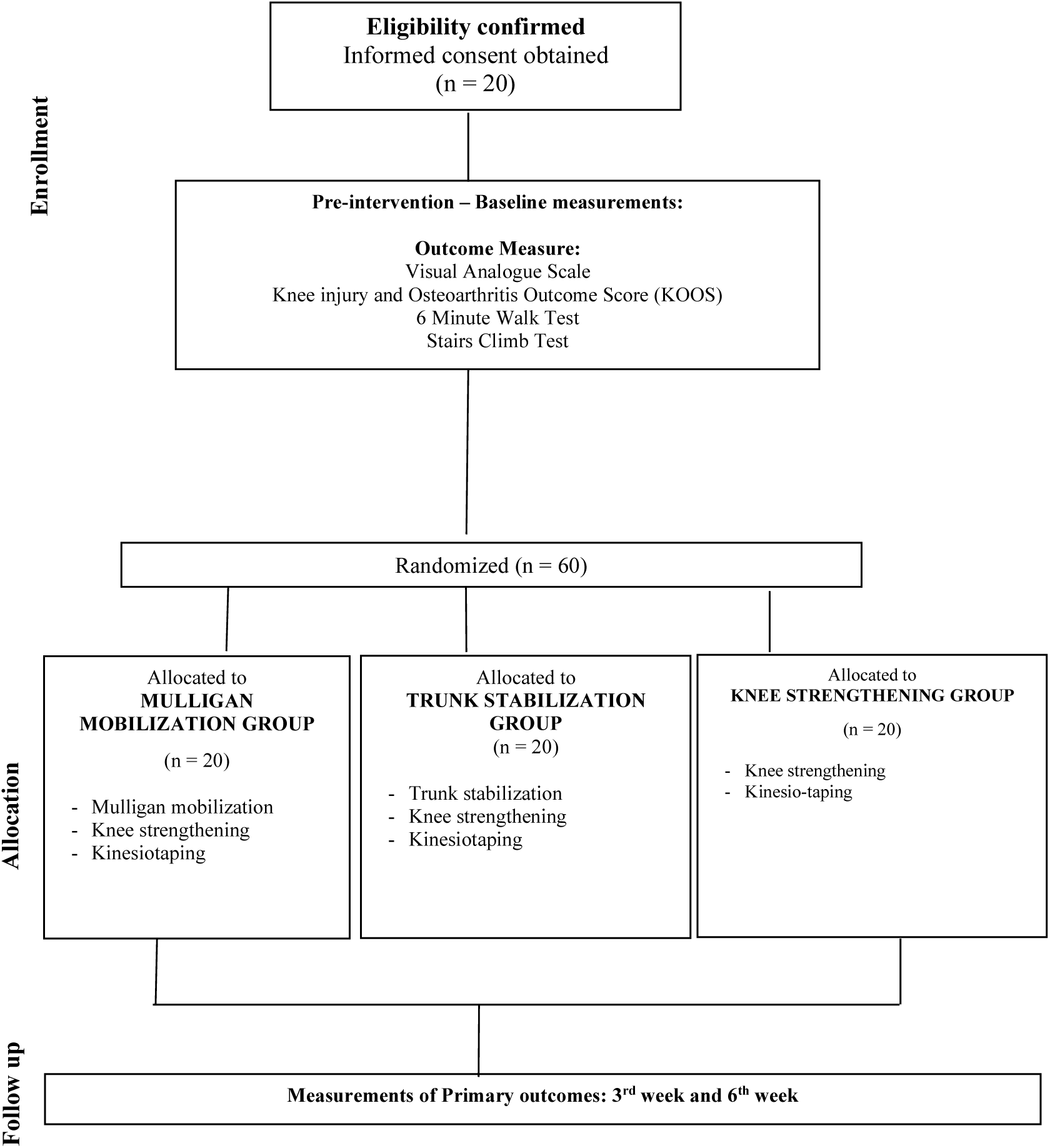
Flow diagram of the planned protocol pathway.

### Mulligan joint mobilization

**Area:** Knee Joint

**Mobilization:** Non-weight bearing (NWB) to weight-bearing (WB)

**Intensity:** 6–10 reps

**Frequency:** 3 sets/ session

**Procedural detail:**

Joint mobilization will be performed in the sagittal, frontal and transverse direction, glide given will depending on patient adherence to joint mobilization. Mobilization will progress from NWB to WB, according to the compliance of the patient. ^24^

### Trunk stabilization

**Area:** Trunk

**Type of exercise:** Stabilization exercise

**Intensity:** 6-8 reps

**Frequency:** 3 sets per session and 30-second duration break between sets

**Procedural detail:**

**i**. Participants will start in the prone position on the bed. They will raise and lower their legs without flexing knees. The lumber region will be stabilized during this exercise to prevent the arms from moving.

**ii. Back bridge:**

The participant will lie supine on the bed with knees flexed at 90 degrees, and lift the pelvis to align the spine and thighs.

**iii. Unilateral back bridge:**

In back-bridge position with the pelvis lifted and a neutral spine position, extend one knee by lifting the foot off the ground.

**iv. lateral step up:**

Standing at the side of a 10-cm step, step up sideways onto to the step. ^25^

### Knee Strengthening

**Area:** Knee Joint

**Type of exercise:** Knee Isometric Strengthening exercises.

**Load:** maximum isometric contraction

**Intensity:** 10 reps

**Frequency:** Beginning with one set of all knee exercises then increasing to two sets at three weeks, and three sets for the remainder of sessions.

**Procedural detail:**

**i. Isometric quadriceps exercise:** The patient will lie supine on the bed. A rolled-up towel will be placed under the knee. They will be instructed to press the knee on the towel to maximally activate the quadriceps isometrically. The participant will hold this position for 5 seconds.

**ii. Straight leg raising (SLR) exercise:** The participant will lie supine on the bed They will be instructed to perform a maximum isometric quadriceps contraction prior to the lifting phase of the exercise. Then they will be instructed to lift the leg up to 10 cm above the plinth and hold the contraction during the lifting phase for 10 seconds. ^26–27^

### Kinesio-taping

The muscle-stretch method will use one Y and two I straps: The Y-shaped strap will be fixed over the top of patella, and the knee will be bent as far as possible before both ends of Y strip are placed around the patella ending on the tibial tuberosity. Reinforcing I-tape will be fixed at the origin and insertion of MCL and LCL, and changed after every session. ^28^

### Measurement of outcomes

In patients with unilateral osteoarthritis, only the affected knee will be examined throughout the study. For patients with bilateral KOA, the most painful knee will be studied. ^29^

### Visual Analogue Scale

This measures subjective pain ranges in 10-cm intervals with defined cut-off scores.^30^ It will be measured at rest and while the patient is ascending and descending stairs. A baseline reading will be taken at the beginning of the study for comparison with further readings at 3^rd^ week and 6^th^ week

### Knee injury and Osteoarthritis Outcome Score (KOOS)

The Knee injury and Osteoarthritis Outcome Score (KOOS) is a patient-reported outcome measure intended for use by middle-aged and elderly patients with knee osteoarthritis (OA) to monitor disease course and outcomes during interventions. KOOS has five subscales and their reliability (1) Pain (r=0.93) (2) other Symptoms (r=0.85) (3) Activities of Daily Living (r=0.95) (4) Sport and Recreation function (r=0.75) and (5) knee-related Quality of Life (r=0.79). ^31^ Each subscale generates a final score ranging from 0 to 100, where 0 represents “worst” and 100 “best”.^32^

### 6-Minute Walk Test

The 6-Min Walk Test (6 MWT) is a test of sub-maximal exercise that entails measuring distance walked over a span of six minutes. Bright-coloured tapes will be used to mark each end of the 12 m walkway. The environment will be free of hazards and readings will be recorded by a assessor who has no knowledge of the group to which the patient belongs. Patients will be instructed to wear comfortable shoes. ^33^

### Stairs Climb Test

The Stair Climb Test is used to measure the total time taken by the participant to ascend and descend a set of stairs with a step height of 16 cm (ICC=0.90). If safety is of concern, the assessor will walk behind the participant going up the stairs and at the side while going down the stairs. If there is no concern for safety, the tester will remain at the start/finish position on the ground landing. ^34^

### Rescue medicine

Any use of paracetamol will be ascertained at weeks 3 and 6 after randomization.

### Adverse effects

There have been no significant risks and no side effects recorded with this treatment intervention apart from a temporary increase in pain, fatigue, muscular strain and skin irritation. In the case of any related event, application of cryotherapy and rest will be considered.

### Data management

The subject will be identified by a serial number rather than by his/her name to ensure total anonymity at all times. All identification will be removed from the data for individual participants before collection. although the principal researcher will have access to participants’ personal data. After completing this study, data will remain the property of the Dow University of Health Sciences. Data will be stored from three months and to three years after article publication.

### Sample size

The sample size was calculated using PASS software. In the pain section, Group1 2.23+0.73, Group2 3.12+0.66, Group3 3.98+0.73 we used this difference to calculate the sample size. We included a power of 99%, confidence interval of 99% and a sample size of 11 per group by using one-way ANOVA. To manage the dropout rate, the sample size is set at 20 per group. ^35^

### Statistical analysis

Data will be entered and analyzed using SPSS version 23. Comparison of group demographics will be conducted using the One-Way Anova analysis for continuous variables, in this case, age, Body Mass Index and duration of symptoms. Chi-square analysis will be used to compare groups using category data such as gender and K/L criteria. Mean and SD will be calculated for quantitative variables such as pain, knee function, sub-maximal exercise capacity and stair climbing using repeated-measure ANOVA. For pairwise comparison, post hoc Tukey will be applied. A value of less than 0.05 will be considered significant. Analysis of Intention to treat will also be carried out in this research.

## Discussion and conclusions

A double-blinded randomized controlled trial has been designed to examine the effects of Mulligan joint mobilization and trunk stabilization exercise on pain, disability and sub-maximal exercise performance in the KOA. A systematic review of the effects of orthopaedic manual therapy (OMT) included 11 randomized controlled trials involving 494 patients. The study shows that OMT combined with strengthening exercises may result in reducing pain and improving physical functions in patients with KOA. However, a clear recommendation for treatment was not possible due to high heterogeneity in the design, lack of information on chronicity of the OA and on whether patellofemoral or tibiofemoral compartment of the knee was affected.^36^ The findings of our study are likely to provide clinically relevant information with reduced bias because only outcomes for the most painful knee will be assessed throughout the study. Both clinical and radiological classification criteria are used in this study in order to correctly diagnose tibiofemoral arthritis.

MWM works on the main potential neurophysiological mechanisms, including changes in central pain processing mechanisms and descending pain inhibitory systems. Furthermore, the movement produced while mobilizing the joint can alter the concentration of inflammatory mediators and lead to the deactivation of nociceptors that are activated by this mechanism.^37^

In a cross-sectional study of 220 patients with OA, 95% of the subjects reported a lack of knee confidence in walking; pain during gait and fear of movement.^38^ Most MWM was carried out in a weight-bearing position, and at the same time patients received painless joint movement feedback. This feedback can modulate psychological features such as fear of movement, which leads to an increase in the walking capacity. Additionally, MWM requires muscle activity in a weight-bearing position, which may result in improved motor performance, allowing the patient to gain long term benefits from a formal exercise program.^39^

Trunk stabilization or core exercises have been used to improve dynamic lower limb alignment and lumbopelvic movement control. The forces acting on the tibio-femoral joint play an important role in knee injuries. Because the location of the body’s centre of mass is largely influenced by the mass of the trunk, aberrant motions of the pelvis and trunk can affect the orientation of the ground force vector and therefore influence the knee load. For instance, weakness in the hip abductor muscle or the external rotator muscle causes the centre of gravity to move to the contralateral pelvis joint. This, in turn, will lead to an increased adjustment of biomechanical forces in the medial compartment of the knee.^40–41^

Our study has some limitations. Participants with medicine prescription will be included in this study. However, research team will maintain medicine prescription record. Other limitation is blinding of the physiotherapist and it is not possible in this study because principle investigator is treating all patients. We are confident, however, that this protocol holds promise, especially once the current drawbacks have been addressed.

## Data Availability

The data that will be generated from this study will not be deposited in a public repository due to privacy and consent restrictions. De-identified data can be made available from the corresponding author on reasonable request, subject to a data sharing agreement.

## Acknowledgement

None

## ICMJE Author roles

SNBN and AS conceived and designed the study. SNBN, SSA and AS were involved in acquisition of data. SNBN and AS drafted this protocol paper. All authors revised and approved the final manuscript.

## Ethics and Dissemination policy

This study has been approved by Institutional Review Board (IRB), Dow University of Health Sciences.

